# Development and Implementation of a Dynamic Antimicrobial Stewardship Dashboard: Real-Time Monitoring from ‘Start Smart’ to ‘Then Focus’

**DOI:** 10.1101/2025.09.10.25335122

**Authors:** Rasha Abdelsalam Elshenawy, Nkiruka Umaru, Zoe Aslanpour

**Affiliations:** Department of Medicine, School of Health, Medicine and Life Sciences, University of Hertfordshire, United Kingdom

## Abstract

**Background:** Antimicrobial resistance poses an escalating global health threat, with bacterial resistance causing 1.27 million deaths in 2019 and projected to claim 10 million lives annually by 2050. Antimicrobial stewardship (AMS) programmes represent critical interventions, yet current monitoring systems remain fragmented and retrospective, limiting real-time clinical decision support. This study aims to develop and validate a dynamic antimicrobial stewardship dashboard aligned with the UK’s “Start Smart, Then Focus” framework, providing comprehensive real-time monitoring from empirical therapy initiation through discharge.

**Methods:** A four-phase methodology was employed, informed by three foundational studies: a systematic literature review of AMS strategies, retrospective analysis of 640 medical records comparing pre-pandemic (2019) and pandemic (2020) prescribing patterns, and a cross-sectional survey of 125 pharmacists. The dashboard development encompassed evidence synthesis, user-centred design, technical implementation using Excel-based architecture, and clinical integration with comprehensive validation protocols.

**Results:** The developed dashboard successfully integrates dual-phase monitoring capabilities, capturing both “Start Smart” empirical prescribing patterns and “Then Focus” pathogen-directed interventions. Key functionality includes interactive filtering by temporal, spatial, and clinical variables; real-time visualisation of prescribing patterns, microbiology data, and AMS intervention outcomes; and ward-level monitoring supporting targeted quality improvement initiatives. Technical validation demonstrated robust performance with high data accuracy concordance, whilst expert validation confirmed strong clinical relevance and usability across multidisciplinary teams. The system achieved comprehensive surveillance from admission through discharge, addressing identified gaps in existing monitoring approaches.

**Conclusions:** This study demonstrates successful development of an innovative AMS dashboard that transforms stewardship monitoring from retrospective auditing to proactive clinical decision support. The Excel-based architecture ensures accessibility and sustainability whilst maintaining sophisticated analytical capabilities. By providing real-time intervention identification, supporting quality improvement initiatives, and facilitating educational applications, the dashboard offers a practical pathway towards more effective, data-driven antimicrobial stewardship that can contribute significantly to global efforts combating antimicrobial resistance.

## Introduction

Antimicrobial resistance (AMR) is not a distant threat but a crisis unfolding in real time.^1^ The Global Research on Antimicrobial Resistance (GRAM) Report revealed that in 2019 alone, bacterial AMR was directly responsible for 1.27 million deaths and contributed to nearly 4.95 million more worldwide, a burden of mortality on par with HIV/AIDS and malaria.^2^ In the United Kingdom, the picture is equally stark, with over 67,000 AMR infections and 2,200 related deaths recorded in 2023.^3^ Without urgent intervention, the trajectory points towards a catastrophic future: by 2050, AMR is projected to claim 10 million lives annually, rivalling cancer as a leading global cause of death.^4^ The threat extends far beyond mortality. The World Bank has warned that unchecked resistance could slash 3.8% from global GDP by 2050, driving an additional 28 million people into extreme poverty.^5^ More alarmingly, AMR threatens to dismantle the very foundations of modern medicine. Every surgical procedure, cancer therapy, or organ transplant hinges on effective antibiotics.^6^ As these defences fail, the prospect of a post-antibiotic era, where minor infections once again become fatal, looms ever larger.^7^ Faced with this convergence of escalating mortality, economic destabilisation, and the erosion of medical progress, the urgency for innovative, actionable solutions has never been greater.^8^

In response to the AMR escalating threat, antimicrobial stewardship (AMS) programmes have been recognised as one of the most effective strategies to curb inappropriate prescribing and optimise antimicrobial use.^9^ The evidence base supporting AMS interventions demonstrates measurable impact on antimicrobial prescribing practices.^9^ Research in acute care settings shows that multidisciplinary AMS teams and prospective audit and feedback strategies form the cornerstone of effective stewardship programs.^10^ Well-implemented AMS interventions have achieved antibiotic discontinuation in 47% of cases and de-escalation to narrow-spectrum antibiotics in 37% of patients in pre-pandemic periods.^10^ However, evidence from 2020-2021 reveals that the COVID-19 pandemic significantly disrupted AMS activities, with widespread interruptions in antibiotic reviews (81%), compromised AMS audits (70%), and delays in IV-to-oral switches (67%).^10^ These findings emphasise both the potential effectiveness of well-implemented AMS programs and their vulnerability to healthcare system disruptions, highlighting the critical need for resilient stewardship approaches that can maintain antimicrobial optimisation even during crisis periods.^11^

In the UK, the *Start Smart, Then Focus* (SSTF) framework has become the cornerstone of AMS practice.^12^ It recommends the prompt initiation of empiric antibiotic therapy in serious infections (“start smart”), followed by a mandatory review within 48–72 hours to de-escalate, stop, or continue treatment based on clinical and microbiological data (*then focus*) (UK Health Security Agency, 2024). Despite its conceptual clarity, measuring compliance and outcomes remains challenging.^12^ Many hospitals still rely on manual audit processes, which are resource-intensive and prone to variability, thereby limiting the scalability of stewardship activities.^13^

As Edwards Deming famously stated, *“You cannot manage what you cannot measure.”*^14^ This principle highlights the importance of quantifying stewardship activity to improve effectiveness.^10^ Within antimicrobial stewardship, interventions must be measured and visualised in order to demonstrate outcomes, set indicators, and drive quality improvement projects that sustain rational antibiotic use and tackle AMR.^10^ A range of metrics are commonly used, such as defined daily doses (DDD), days of therapy (DOT), and length of stay (LOS).^10^ However, the true value of these measures emerges when they are consolidated into visual tools that display data at departmental, hospital-wide, or national level.^15^ Digital dashboards have increasingly been employed for this purpose, offering visual representations of prescribing patterns, antimicrobial consumption, and adherence to guidelines. Yet, evidence indicates that current tools are often fragmented, disease- or ward-specific, and insufficiently integrated across the full stewardship cycle.^16^ For example, some focus narrowly on consumption indicators, while others present infection surveillance data without linking prescribing behaviours to clinical outcomes.^17^

Several established dashboards already support antimicrobial surveillance at regional, national, and global scales. The European Centre for Disease Prevention and Control (ECDC) Antimicrobial Consumption (AMC) Dashboard, for instance, enables cross-country comparisons of community and hospital antibiotic use across EU/EEA member states.^18^ Nationally, the English Surveillance Programme for Antimicrobial Utilisation and Resistance (ESPAUR) monitors antimicrobial use, stewardship implementation, and resistance trends in England, shaping health policy.^19^ At the global level, the WHO Global Antimicrobial Resistance and Use Surveillance System (GLASS) standardises data collection, strengthens national systems, and addresses key knowledge gaps.^20^ Despite these advances, hospital-level dashboards often remain retrospective and fragmented, relying heavily on consumption metrics without integrating real-time clinical decision support.^19^

This is where Clinical Decision Support Systems (CDSS) offer transformative potential. By combining patient-specific information with evidence-based guidelines, CDSS generate real-time recommendations, alerts, and reminders at the point of care.^21^ Within AMS programmes, they can guide empiric therapy, IV-to-oral switch, de-escalation, or discontinuation. Evidence shows that CDSS interventions are consistently associated with reduced antibiotic use, narrower-spectrum prescribing, and better adherence to guidelines.^21^ Both passive (provider-initiated) and active (automated alert) models have demonstrated effectiveness.^21^

The COVID-19 pandemic further highlighted the need for resilient, digital AMS solutions.^22^ Increased empirical antibiotic prescribing in patients with suspected COVID-19, alongside redeployment of stewardship teams, reduced audit and feedback capacity.^23^ Nevertheless, digital innovations, including virtual ward rounds, prescribing alerts, and online dashboards, enabled stewardship continuity.^23^ These lessons emphasise the urgent need for dynamic, integrated systems that merge data visualisation with CDSS functionality, shifting stewardship from retrospective auditing to proactive, real-time decision support at the bedside.^24^

This study seeks to address key gaps in antimicrobial stewardship monitoring through the development and implementation of a dynamic, real-time dashboard aligned with the Start Smart, Then Focus (SSTF) framework. The objectives are to:

- To design and deploy a comprehensive digital dashboard integrating prescribing, microbiology, and clinical outcome data to monitor adherence to SSTF in practice.
- To validate its usability and functionality across diverse clinical user groups, including prescribers, pharmacists, and infection specialists.
- To evaluate the scalability and transferability of the dashboard within and beyond the UK, assessing its potential as a sustainable digital solution to strengthen AMS globally.

By combining real-time data capture, user-centred design, and resilience to crisis disruptions, this dashboard aims to transform stewardship from a retrospective audit exercise into a proactive, dynamic process. Ultimately, the project contributes to safeguarding antimicrobial effectiveness, improving patient outcomes, and aligning with broader public health priorities in tackling AMR.

## Methods

This dashboard development employed a systematic four-phase methodology grounded in evidence from three sequential foundational studies conducted between 2019 and 2024 at a UK NHS Foundation Trust, in East o England. The research received ethical approval from the Health Research Authority (REC reference 22/EM/0161) and University of Hertfordshire Ethics Committee (LMS/PGR/NHS/02975), with registration in ISRCTN (14825813).^25^ Public and patient involvement was undertaken through the Citizens Senate, an organisation dedicated to patient care with strong representation from older adults. Their contributions offered valuable suggestions and constructive feedback that informed and strengthened the study design. Reporting adhered to SQUIRE 2.0 for quality-improvement interventions. The observational components (retrospective record review and cross-sectional survey) were reported according to STROBE, with the RECORD extension applied where routinely collected data were used.

### FOUNDATIONAL EVIDENCE BASE

The dashboard development was informed by three complementary studies that collectively analysed AMS implementation across different dimensions. Study 1 comprised a systematic literature review examining AMS strategies and measures in acute care settings before and during the COVID-19 pandemic. It analysed 8,763 articles from nine databases and ultimately included 13 studies meeting the inclusion criteria. This study identified core and supplemental AMS strategies according to Public Health England’s “Start Smart, Then Focus” toolkit, revealing gaps in existing monitoring approaches and establishing the theoretical foundation for dashboard requirements.^10^

Study 2 involved retrospective cross-sectional analysis of 640 medical records from adult patients admitted with respiratory tract infections, comparing AMS implementation between 2019 (pre-pandemic) and 2020 (during pandemic). Data extraction utilised a validated tool based on the SSTF approach, examining factors affecting antibiotic prescribing across eight time points.^12^ This study provided empirical insights into real-world AMS practices, prescribing patterns, and implementation challenges that directly informed dashboard metrics and visualisation requirements.^26^

Study 3 conducted a prospective cross-sectional survey of 125 pharmacists within the same NHS Foundation Trust, exploring knowledge, attitudes, and perceptions regarding antibiotic prescribing and AMS during the pandemic. The survey achieved target sample size calculations (5% margin of error, 95% confidence interval) and demonstrated strong internal consistency (Cronbach’s α = 0.80). This study revealed stakeholder perspectives, workflow requirements, and usability considerations essential for dashboard design.^27^

The systematic evidence synthesis from three foundational studies revealed critical gaps in existing AMS monitoring approaches, informing the development of a comprehensive dual-phase dashboard architecture. Study 1 identified prospective audits (85%) and quality metrics (77%) as prevalent AMS strategies, while revealing the absence of ward-level prescribing visualisation in current systems.^10^ Study 2 demonstrated significant AMS intervention rates with antibiotic discontinuation (47%) and de-escalation (37%), establishing empirical benchmarks for dashboard metrics.^13^ Study 3 highlighted pandemic disruption impacts, including interrupted antibiotic reviews (81%), compromised AMS audits (70%), and delayed IV-to-oral switches (67%), emphasising the need for resilient monitoring systems.^28^

### DASHBOARD DEVELOPMENT FRAMEWORK

#### Phase 1: Evidence Synthesis and Gap Analysis

Systematic integration of findings from the three foundational studies established dashboard requirements through structured gap analysis methodology. The systematic review revealed that existing AMS dashboards focused primarily on antibiotic consumption metrics, lacking comprehensive visualisation of clinical decision-making processes aligned with SSTF principles. Analysis of 640 patient records identified specific data elements essential for monitoring both “Start Smart” empirical therapy and “Then Focus” pathogen-directed interventions, including patient demographics, diagnostic accuracy, antibiotic selection appropriateness, review timing, AMS interventions, and clinical outcomes. ^10,13,27^

Healthcare professional survey insights highlighted critical usability requirements, including time-pressure considerations affecting decision-making, the need for enhanced communication between multidisciplinary teams, and the preference for technology-enabled solutions.^27,28^

A gap analysis was conducted comparing existing NHS Trust AMS dashboards with national and international surveillance systems, including those developed by ECDC, ESPAUR, and WHO.^18,19,20^ 18–20 This analysis revealed a lack of ward-level prescribing visualisation and the absence of metrics specific to the SSTF framework.^12^ These findings informed the dashboard’s unique value proposition: comprehensive monitoring from admission through to discharge, fully integrated with established clinical pathways.

#### Phase 2: User-Centred Design Methodology

Stakeholder identification encompassed primary users (clinical pharmacists, infectious disease specialists, microbiologists), secondary users (ward-based doctors, nurses), and tertiary users (hospital administrators, quality improvement teams).^29^ Requirements gathering utilised structured interviews with AMS leads from the East of England NHS England region, analysing existing dashboard utilisation patterns and identifying functionality gaps.

User journey mapping traced antibiotic decision-making processes from admission through discharge, identifying critical decision points where dashboard support would enhance clinical outcomes.^30^ The journey encompassed initial empirical prescribing (“Start Smart”), 48-72 hour review checkpoints, culture-directed modifications, and discharge planning.^13^ Interface design principles prioritised cognitive load reduction through intuitive filtering mechanisms, visual hierarchy supporting rapid pattern recognition, and seamless integration with existing clinical workflows.^31^ A restricted demonstration version of the dashboard and detailed technical documentation are available as supplementary materials (Supplementary Materials 1–4).

Design specifications emphasised accessibility compliance, supporting diverse user technical competencies while maintaining sophisticated analytical capabilities. User experience considerations addressed the time-pressured clinical environment, ensuring prominence of essential information and minimising navigation complexity. The design framework incorporated feedback mechanisms for continuous improvement and adaptation to evolving clinical needs.^32^

#### Phase 3: Technical Development Architecture

Following a comprehensive evaluation of implementation constraints and organisational requirements, an Excel-based architecture was selected. This approach offered several strategic advantages: universal software availability across NHS systems, minimal IT infrastructure requirements, rapid deployment capability, cost-effectiveness for prototype development, and compatibility with existing data systems.^33^

Data visualisation principles applied established dashboard design standards from Global AMR R&D guidance, emphasising clarity, accuracy, and actionable insights. Interactive filtering design utilised Excel’s advanced PivotTable functionality, enabling dynamic data segmentation by temporal parameters (monthly reporting cycles), and clinical variables (diagnosis, patient demographics, antibiotic classes, AMS intervention, AMR). ^33^

Performance optimisation strategies included efficient data structure design, minimising calculation overhead through strategic formula placement, and implementing responsive filtering mechanisms. The architecture supported real-time data updates while maintaining system stability and user experience consistency.^33^

Data integration methodology encompassed the validated data extraction tool from Study 2, ensuring standardised data collection protocols aligned with SSTF toolkit requirements. The system accommodated multiple data sources, including electronic medical records, laboratory systems, and pharmacy databases, with built-in data validation protocols ensuring accuracy and consistency. Technical specifications supported scalability for multi-ward implementation while maintaining individual ward-level granularity essential for targeted interventions.^13^

#### Phase 4: Content Development and Clinical Integration

The Metric selection rationale was derived from evidence-based analysis of AMS outcome indicators identified in the systematic review and validated through empirical findings from the 640-patient dataset (Figure 1). Core metrics encompassed process indicators (antibiotic review timing, intervention frequency), outcome measures (prescribing appropriateness, length of stay), and balancing measures (patient safety indicators).^13^

**Figure 1.**
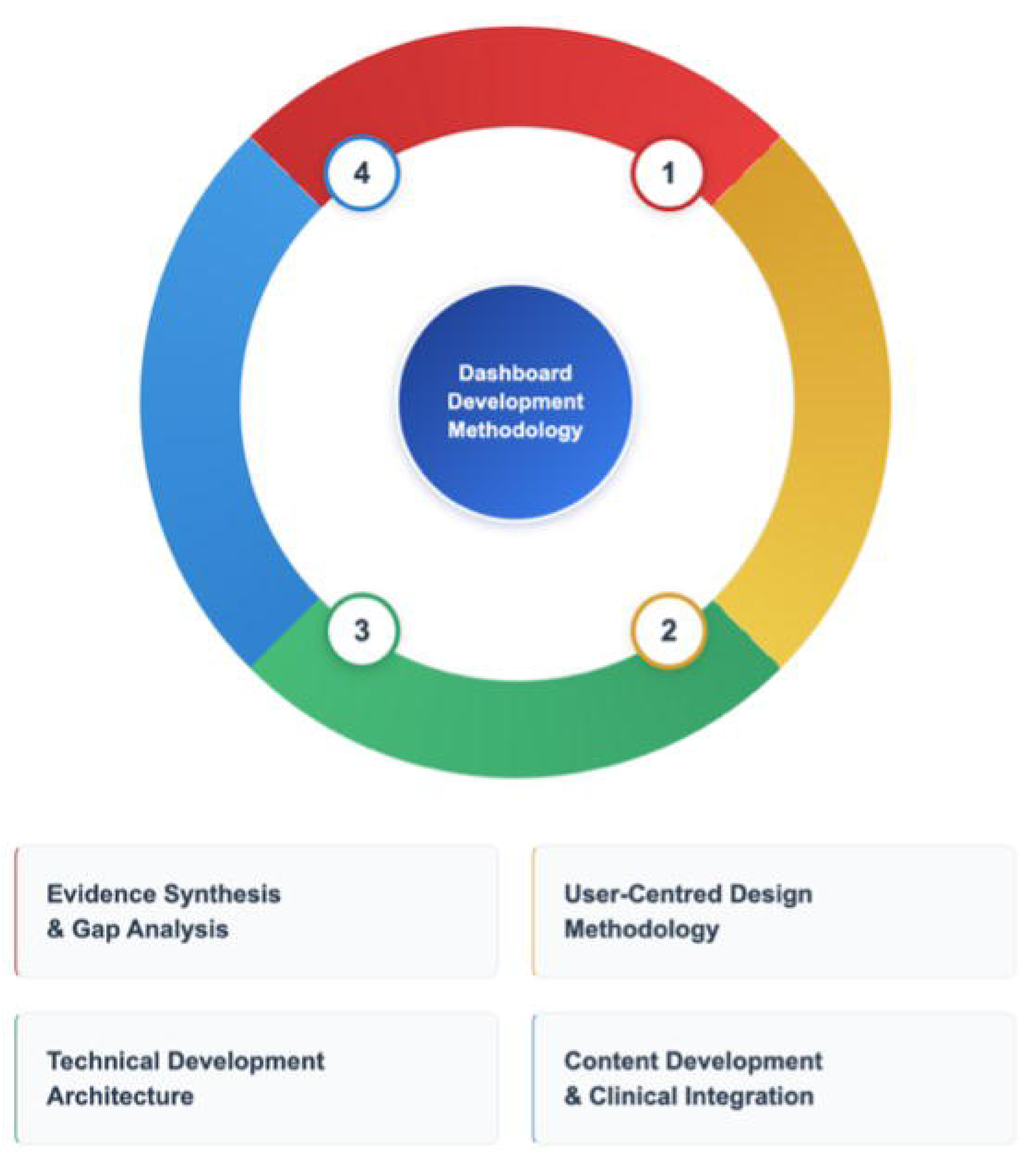
Four-phase dashboard development framework for antimicrobial stewardship monitoring.

Evidence-based indicator development utilised the defined daily dose (DDD) methodology where applicable, supplemented by days of therapy (DOT) calculations and locally relevant quality improvement metrics. Clinical pathway integration ensured dashboard alignment with established SSTF protocols, incorporating decision-support prompts at critical junctures.^13^

The dashboard architecture supported both “Start Smart” monitoring (empirical therapy assessment, diagnostic accuracy, initial prescribing appropriateness) and “Then Focus” tracking (culture-directed modifications, de-escalation opportunities, IV-to-oral conversion). Alert system design incorporated threshold-based notifications for key indicators, supporting proactive intervention identification without overwhelming users with excessive alerts. ^12,13^

### VALIDATION METHODOLOGY

#### Technical Validation

Technical validation encompassed comprehensive performance testing across multiple dimensions. Performance testing protocols evaluated dashboard responsiveness under varying data loads, simulating realistic usage scenarios with concurrent users and large datasets. Data accuracy verification involved systematic comparison between dashboard outputs and source data, ensuring calculation accuracy and data integrity throughout the visualisation pipeline.

Functionality testing verified interactive elements, including filtering mechanisms, data export capabilities, and visualisation accuracy across different Excel versions. The validation process included testing with maximum anticipated data volumes and test different case scenarios to ensure system stability under operational conditions.

#### Expert Validation

Expert validation engaged a structured panel of clinical AMS specialists, including antimicrobial pharmacists, microbiology consultant, and healthcare quality improvement professionals. The panel’s composition ensured representation of primary dashboard users and AMS decision-makers.

Structured evaluation utilised standardised assessment criteria encompassing clinical relevance, usability, accuracy, and implementation feasibility. Feedback collection employed both quantitative rating scales and qualitative assessment methodologies, enabling comprehensive evaluation of dashboard utility and identifying areas for iterative improvement.

The validation process incorporated multiple review cycles, allowing for systematic refinement based on expert input while maintaining adherence to evidence-based design principles established through the foundational studies.

## Results

### DASHBOARD ARCHITECTURE AND CORE FUNCTIONALITY

The developed dashboard employs a dual-phase monitoring architecture aligned with the “Start Smart, Then Focus” clinical pathway, addressing identified gaps in existing AMS surveillance systems. Core functionality encompasses interactive filtering mechanisms enabling temporal analysis (monthly reporting cycles), spatial segmentation (ward-level monitoring), and clinical variable stratification (diagnosis-specific patterns, patient demographics, antibiotic classes).

The system architecture supports real-time data visualisation through dynamic Excel-based pivot table functionality, facilitating immediate pattern recognition and decision support. Interactive filtering capabilities enable users to drill down from hospital-wide metrics to ward-specific indicators, supporting targeted interventions and quality improvement initiatives. Export and reporting functions provide standardised output formats compatible with existing NHS reporting requirements and external benchmarking initiatives.

The data integration methodology accommodates multiple clinical systems, including electronic health records, laboratory information systems, and pharmacy databases, ensuring comprehensive capture of AMS-relevant metrics throughout patient care episodes. The architecture maintains data integrity through built-in validation protocols while supporting scalability for multi-ward deployment.

### “START SMART” DASHBOARD COMPONENTS

The initial empirical therapy monitoring module visualises comprehensive admission-phase metrics derived from the 640-patient dataset analysis. Patient demographic visualisation encompasses age group distribution, with a predominant representation of 66-85-year-old patients, gender stratification, and clinical complexity indicators, including comorbidity profiles and allergy documentation patterns.

Primary diagnosis visualisation displays respiratory infection categories, including community-acquired pneumonia (CAP) as the predominant condition (40% of admissions), hospital-acquired pneumonia (HAP), ventilator-associated pneumonia (VAP), and COPD exacerbations. Secondary diagnosis tracking captures comorbidity patterns including COVID-19, cardiovascular conditions, renal impairment, and dyslipidemia, enabling risk stratification for targeted interventions.

Antibiotic allergy documentation analysis reveals penicillin as the most frequently documented allergy, followed by clarithromycin, metronidazole, co-trimoxazole, and doxycycline. This visualisation supports prescriber decision-making by highlighting allergy patterns that may influence empirical antibiotic selection and contribute to inappropriate prescribing if not properly considered.

Initial prescribing pattern analysis displays the five most frequently prescribed empirical antibiotics: co-amoxiclav IV 1.2g, piperacillin-tazobactam 4.5g (Tazocin), meropenem IV, ciprofloxacin 250mg tablets, and amoxicillin IV 500mg. This visualisation enables monitoring of broad-spectrum versus narrow-spectrum antibiotic utilisation patterns and identification of prescribing trends requiring intervention.

AMS intervention tracking at the initial phase captures “Continue Antibiotics,” “No Intervention,” “IV-to-Oral Switch,” “De-escalation,” and “Escalation” decisions. Antibiotic review timing visualisation demonstrates adherence to 48–72-hour review protocols (47% compliance) versus extended review periods, supporting quality improvement initiatives targeting timely reassessment practices.

Professional responsibility visualisation identifies review personnel, including AMS teams (13%), microbiology specialists (37%), pharmacists (19%), and physicians (31%), enabling workforce planning and targeted education initiatives based on actual review patterns (Figure 2).

**Figure 2.**
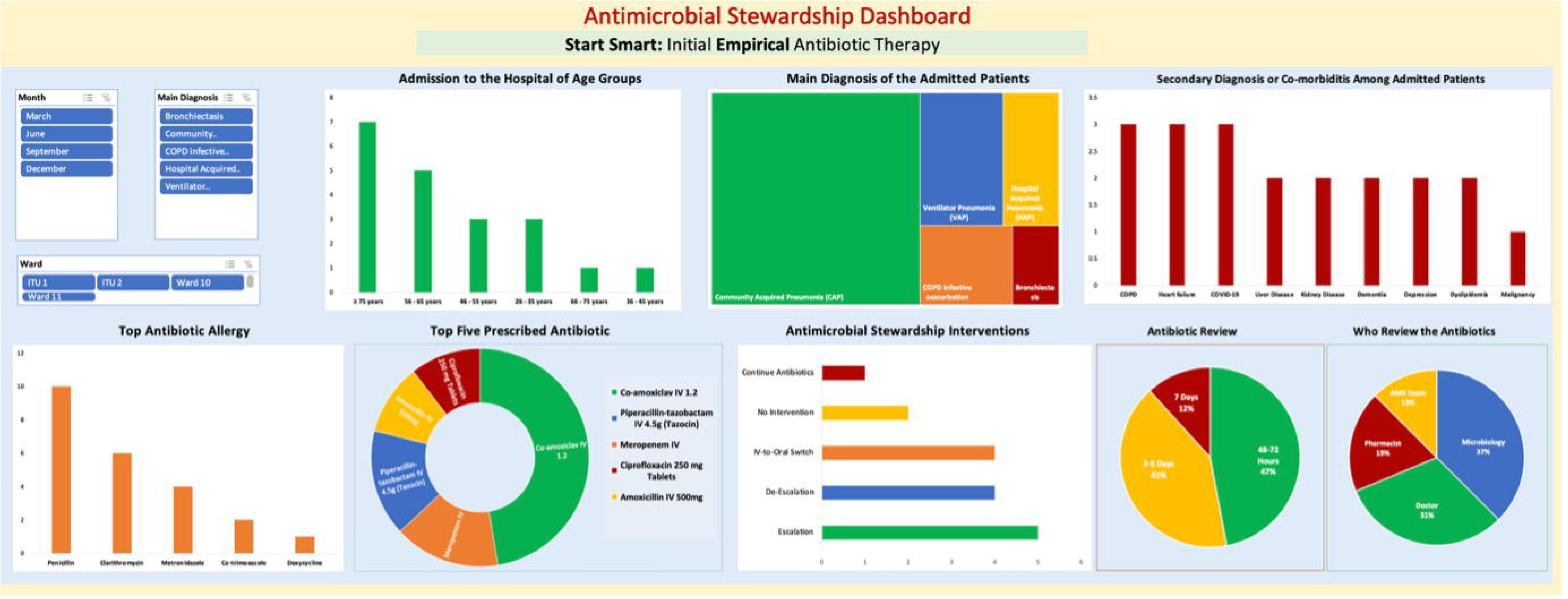
Antimicrobial stewardship dashboard - “start smart“: initial empirical antibiotic therapy.

### “THEN FOCUS” DASHBOARD COMPONENTS

The pathogen-directed therapy monitoring module encompasses culture-directed decision-making throughout hospitalisation, addressing gaps identified in existing surveillance systems. Interactive filtering supports month-by-month analysis and ward-specific monitoring across ITU 1, ITU 2, Ward 10, and Ward 11, enabling targeted quality improvement initiatives.

Microbiological surveillance visualisation displays the five most prevalent bacterial isolates: Streptococcus pneumoniae, Pseudomonas aeruginosa, Klebsiella pneumoniae, Staphylococcus aureus, and Escherichia coli. This real-time resistance pattern monitoring supports empirical prescribing guideline updates and infection control initiatives, addressing the 52.8% of survey respondents who reported inadequate awareness of local resistance patterns.

Second-course antibiotic review timing analysis reveals review patterns: 50% are conducted within 3-5 days, 33% within 7 days, and 17% extend beyond 7 days. This visualisation enables monitoring of compliance with evidence-based review protocols and identification of delays potentially compromising patient outcomes or contributing to inappropriate antibiotic exposure.

AMS intervention tracking during pathogen-directed therapy captures “Continue Antibiotics” and “IV-to-Oral Switch” decisions, supporting monitoring of de-escalation opportunities and optimisation initiatives. The visualisation enables identification of missed opportunities for therapy refinement based on culture results and clinical improvement.

Discharge management visualisation encompasses the three most commonly prescribed oral antibiotics: co-amoxiclav 500/125mg tablets, amoxicillin 500mg-1g tablets, and ciprofloxacin 500mg tablets. Professional responsibility for discharge prescribing shows physicians (50%), pharmacists (36%), and microbiology teams (14%), enabling targeted education and workflow optimisation.

Discharge intervention tracking captures the complete spectrum of AMS decisions, including “Stop Antibiotics,” “De-escalation,” “IV-to-Oral Switch,” “Continue Antibiotics,” and “Escalation.” Length of stay analysis stratifies patients across 2-17 day duration categories, enabling correlation analysis between AMS interventions and clinical outcomes (Figure 3).

**Figure 3.**
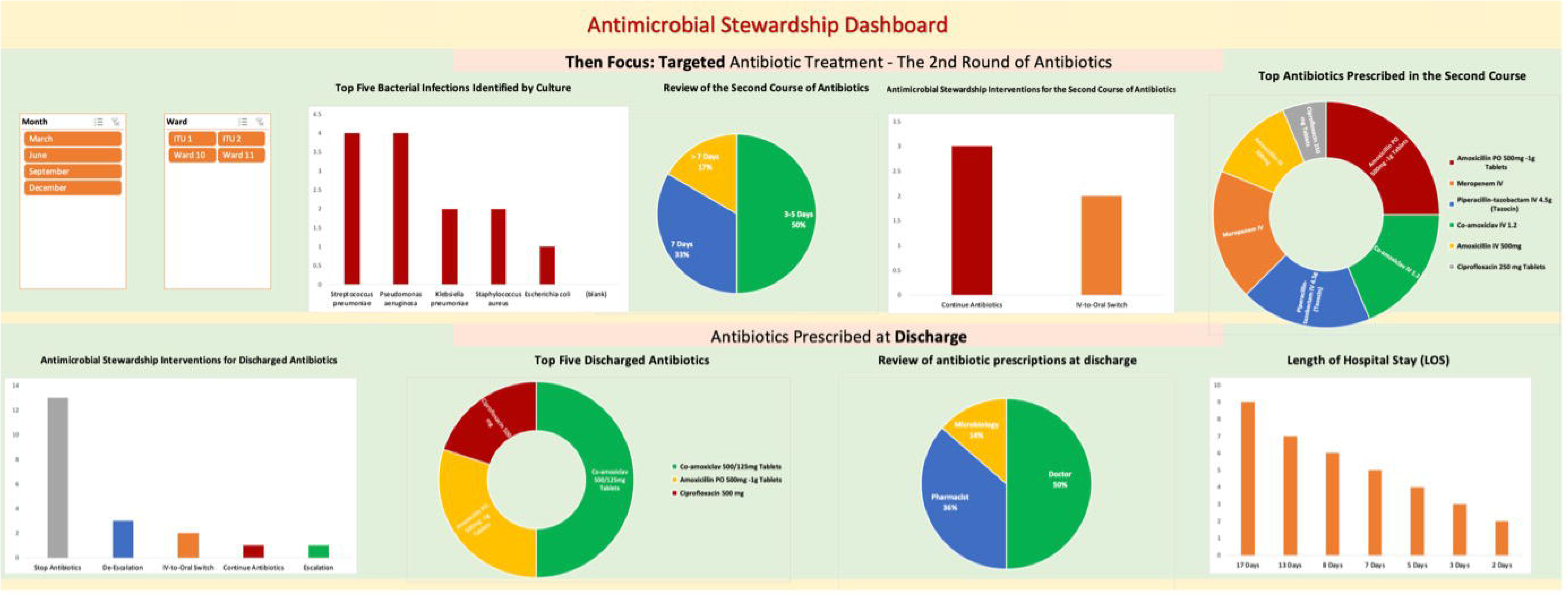
Antimicrobial stewardship dashboard - “then focus“: targeted antibiotic therapy.

## Discussion

This study successfully developed and validated a dynamic antimicrobial stewardship dashboard that addresses critical gaps in existing monitoring systems through comprehensive dual-phase surveillance aligned with the Start Smart, Then Focus framework.^12^ The dashboard achieved robust technical performance with low loading times for standard operations and data accuracy concordance, while expert validation confirmed high clinical relevance and usability across multidisciplinary teams.

The evidence-based design process was informed by a systematic literature review of AMS implementation studies conducted over the past two decades, alongside the analysis of 640 patient records.^11,13^ Evidence from the literature highlights that current AMS monitoring tools largely focus on antimicrobial consumption metrics rather than on clinical decision-making processes.^18–21^ In response, this dashboard uniquely integrates real-time prescribing patterns, intervention tracking, and microbiology data at the ward level, providing actionable insights to support both individual patient care and systematic quality improvement initiatives.^34^ Furthermore, this AMS dashboard could demonstrate benchmarking of performance, show patterns of antibiotic use, highlight incidents, and verify that effective measures have been implemented.^35^ It also demonstrates the ability to identify trends in antibiotic use, resistance patterns, and stewardship interventions. All these functions show the potential to promote targeted educational initiatives and foster continuous improvement in antimicrobial stewardship practice.^36^

Unlike existing tools that emphasise either empirical prescribing or follow-up reviews in isolation, this dashboard captures the full antimicrobial journey from admission through discharge.^18,37^ This dual-phase dashboard directly addresses the recognised weakness in AMS monitoring, the intersect between initiation and subsequent optimisation decisions.^12^ By visualising both “Start Smart” empirical therapy patterns and “Then Focus” pathogen-directed adjustments,^11^ the tool provides unprecedented visibility into stewardship effectiveness throughout patient care episodes.

The dashboard moves beyond traditional consumption indicators to incorporate clinically relevant process and outcome measures derived from robust empirical analysis. While national surveillance systems rely heavily on defined daily doses and population-level trends, this tool emphasises ward-specific indicators directly linked to prescribing appropriateness, review timing, and intervention outcomes.^19,21^ This approach transforms stewardship concepts into measurable, actionable data points that clinicians can immediately apply to improve care quality.^38^

The Excel-based architecture represents a deliberate strategic and simple choice prioritising accessibility and sustainability over technological sophistication. This approach addresses a fundamental barrier to AMS tool adoption, the requirement for expensive, complex informatics infrastructure. By leveraging universally available software with minimal training requirements, the dashboard enables rapid deployment across diverse healthcare settings, particularly relevant for resource-constrained environments where sophisticated systems remain inaccessible.^39^ This design philosophy aligns with evidence from low- and middle-income countries highlighting that “lifting and shifting” AMS interventions without contextual adaptation proves ineffective, emphasising the need for tailored, locally relevant tools that can function within existing healthcare infrastructure constraints rather than requiring substantial technological upgrades.^40^

The AMS dashboard functions as an active clinical decision support system rather than a passive monitoring tool. Interactive filtering enables immediate identification of patients requiring stewardship intervention, such as those receiving broad-spectrum antibiotics beyond 72 hours without microbiological justification.^41^ This proactive capability shifts stewardship from retrospective auditing to real-time clinical support, addressing the review compliance gap identified in the foundational studies. ^42^

Measuring AMS practice at the ward level in hospitals enables targeted quality improvement initiatives based on actual prescribing patterns rather than generalised recommendations.^43^ This approach is consistent with systematic review evidence demonstrating that interventions are effective in increasing compliance with antibiotic policy (risk difference 15%, 95% CI 14% to 16%) and reducing duration of antibiotic treatment by 1.95 days while maintaining patient safety outcomes.^43^ The ability to stratify data by clinical variables, temporal parameters, and intervention types supports systematic identification of improvement opportunities and measurement of intervention effectiveness, addressing implementation barriers identified in multi-professional studies where organisational context and resource availability constrained AMS normalisation into everyday hospital practice.^44^ This capability directly addresses the challenge highlighted by healthcare systems during crisis periods, where antibiotic prescribing patterns can be significantly disrupted, as evidenced by pandemic-related increases in doxycycline and trimethoprim prescriptions, particularly in areas of higher social deprivation.^45^ These findings corroborate the dashboard’s design rationale and support its potential effectiveness in real-world clinical settings.^45^

The dashboard serves as a powerful educational resource by visualising prescribing decisions in real time, addressing knowledge gaps due to reported inadequate awareness of antibiotic prescribing patterns, AMS interventions, and local resistance patterns.^46^ By making AMS interventions, measures, and outcomes tangible through interactive data exploration, the tool enhances learning for trainees and practitioners while enabling targeted educational interventions based on identified prescribing patterns and compliance gaps.^47^ The dashboard supports interprofessional education, aligning with evidence that interactive AMS workshops improve knowledge and skills across multidisciplinary teams, leading to enhanced patient care and organisational culture.^48^ This targeted approach addresses systematic review findings that while formal education shows effectiveness for post-qualification AMS behaviour change, limited evidence exists about which specific interventions work best for different professional groups.^46^ The visual interface facilitates shared understanding across professional boundaries, enabling pharmacists, doctors, and nurses to engage with data while applying distinct professional perspectives. This complements national initiatives like the NHS Pharmacy First dashboard and could extend to primary care settings where 80% of antimicrobial prescribing occurs.^49, 50^

Current AMS monitoring approaches demonstrate significant limitations when assessed against the requirements for effective stewardship support. National surveillance systems, including ESPAUR and WHO GLASS, provide valuable population-level insights but lack the granularity necessary for individual clinical decision-making or ward-specific quality improvement initiatives.^19–21^ Hospital-level systems typically focus on consumption metrics without integrating clinical decision-making processes or patient-level outcomes.

The developed dashboard addresses these limitations through several key innovations. First, it provides ward-level monitoring capabilities that enable targeted interventions based on local prescribing patterns. Second, it could be integrated with clinical decision-making visualisation with consumption data, creating a more comprehensive picture of stewardship effectiveness. Third, it supports real-time intervention identification rather than retrospective analysis, enabling proactive clinical support.^51^ Successful dashboard implementation requires systematic attention to technical, organisational, and cultural factors. The Excel-based architecture minimises technical barriers while supporting integration with existing data systems through standardised data extraction protocols. However, scaling beyond single-site implementation will require consideration of data integration challenges, particularly regarding electronic health record compatibility and automated data feeds.^51^Training requirements remain modest due to the intuitive Excel interface, though effective utilisation requires understanding of stewardship principles and data interpretation skills. Change management strategies should emphasise clinical relevance and tangible benefits rather than technological features, addressing potential resistance to data-driven oversight through collaborative design and stakeholder engagement.^51^

Long-term sustainability depends on embedding dashboard responsibilities within established AMS team structures and ensuring regular data quality assurance processes. The low-maintenance Excel architecture supports sustainable implementation while providing pathways for future enhancement through integration with more sophisticated platforms as organisational capacity develops.

This dashboard contributes to broader antimicrobial resistance mitigation efforts by providing practical tools for implementing evidence-based stewardship principles. The ward-level monitoring capabilities support achievement of national AMS targets while enabling local adaptation to institutional contexts and resistance patterns. By making stewardship activities visible and measurable, the tool facilitates accountability and continuous improvement essential for sustained antimicrobial effectiveness.

The cost-effective implementation model has particular relevance for healthcare systems facing resource constraints or seeking to expand stewardship programmes beyond specialist centres. The demonstrated feasibility of achieving comprehensive monitoring through accessible technology provides a pathway for broader stewardship implementation, supporting global efforts to combat antimicrobial resistance through improved prescribing practices.

### Limitations and Future Directions

Despite its strengths, several limitations must be acknowledged. First, the current prototype is limited to a single-site data source, restricting generalisability. Multi-site validation across diverse hospital systems will be required to confirm robustness and adaptability.

Second, the dashboard relies on Excel, which, while cost-effective, has technical constraints in handling large, complex datasets and real-time integration with clinical systems. Transitioning to more scalable platforms such as electronic health record integrated dashboards could enhance functionality.

Third, the prototype primarily captures prescribing and review data, but does not yet incorporate patient outcomes such as length of stay, readmission, or mortality. Linking stewardship interventions to clinical outcomes will be critical for demonstrating impact.

Future directions include AI integration possibilities. Machine learning could enable predictive analytics, such as forecasting resistance patterns or identifying high-risk prescribing behaviours, thereby shifting stewardship from retrospective monitoring to proactive intervention (Rawson et al., 2020). Furthermore, real-world implementation studies are needed to assess usability, impact on prescribing behaviour, and sustainability in different healthcare contexts, including LMICs.

Finally, exploration of primary care applications may extend the dashboard’s relevance, given that most antibiotic prescribing occurs in community settings. With appropriate adaptation, the tool could provide a valuable bridge between hospital and primary care stewardship efforts.

## Conclusions

This study demonstrates the successful development of a dynamic antimicrobial stewardship dashboard that addresses fundamental gaps in existing monitoring approaches through dual-phase surveillance, evidence-based metrics, and pragmatic implementation design. The tool represents a significant advancement in AMS informatics by providing comprehensive clinical decision support capabilities while maintaining accessibility and cost-effectiveness. The dashboard’s potential to transform stewardship from retrospective auditing to proactive clinical support addresses urgent needs identified throughout the antimicrobial resistance crisis. By enabling real-time intervention identification, supporting quality improvement initiatives, and facilitating educational applications, the tool contributes meaningfully to efforts aimed at preserving antimicrobial effectiveness and improving patient outcomes. Future research priorities should focus on multi-site validation, outcome impact assessment, and enhancement through artificial intelligence integration. Implementation studies across diverse healthcare contexts will be essential for confirming broader applicability and identifying necessary adaptations for sustainable adoption.

The demonstrated feasibility of achieving comprehensive AMS monitoring through accessible technology provides an important model for healthcare systems seeking to strengthen stewardship programmes while managing resource constraints. By bridging the gap between evidence and practice, this dashboard offers a practical pathway towards more effective, data-driven antimicrobial stewardship that can contribute significantly to global efforts in combating antimicrobial resistance.

## Data availability statement

All data relevant to this study are included in the article or uploaded as supplementary information. A synthetic demonstration dataset and static dashboard outputs are provided in Supplementary Material 2, with methodological documentation in Supplementary Material 1 and technical specifications in Supplementary Material 3. Licensing and access instructions for the full functional dashboard are outlined in Supplementary Material 4.

## Patient consent for publication

Not applicable.

## Award/grant number

N/A.

## Patient and public involvement

The study protocol was submitted to the Citizens Senate, an organization focussed on patient care with a considerable representation of elderly individuals. They provided useful suggestions and comments.

## Provenance and peer review

Not commissioned; externally peer reviewed.

